# Conditioning chemotherapy exposure is associated with epigenetic modifications in *Clostridioides difficile* isolates from stem cell transplant recipients

**DOI:** 10.64898/2026.01.12.25342741

**Authors:** Jacob Ng, Jennifer Trannguyen, Rachel Wilkinson, Fritz Conrad, Sam Fehrenbach, Bailey Ebersole, Deb Ghosh, Senu Apewokin

## Abstract

*Clostridioides difficile* is a highly methylated organism within the gut microbiome that is responsible for *Clostridioides difficile* infection (CDI), a common disease that is mediated by toxins production from the bacterium. *C. difficile* infection is ten times more common in chemotherapy patients than the average patient, but the reasons for this disparity are unclear. Conditioning chemotherapy (CC), an integral part of cancer treatments, has the ability to induce methylation changes in many cell types. We posit that CC induces methylation changes within *C. difficile* that may promote toxin production and consequently CDI. To test our hypothesis, we sought to identify the epigenetic changes, particularly methylation changes, within *C. difficile* isolates before and after chemotherapy and within isolates that express toxin and isolates that do not. After stool sampling, we isolated *C. difficile* by culture then sequenced and created a hybrid assembly of each isolate using nanopore long read sequencing and Illumina short read sequencing. Bioinformatics tools such as Dorado and Samtools were used to basecall and determine methylation states, while Unicycler was used for genome assembly. Methylartist was then used for data visualization. Genome-wide methylation profiling revealed distinct epigenetic signatures in *Clostridioides difficile* associated with toxin expression and chemotherapy exposure. Whole-genome 6mA analysis demonstrated significant differences between toxin-positive and toxin-negative isolates, with prominent methylation changes in *tcdA* and *tcdE*, while selected sporulation genes were unmethylated in toxin-negative strains. Chemotherapy was associated with a significant shift in global 6mA methylation patterns. Targeted 5mC analysis of the pathogenicity locus revealed reduced methylation around *tcdB* and across multiple toxin genes following chemotherapy, whereas sporulation genes remained unaffected. These findings indicate chemotherapy-associated epigenetic remodeling of toxin-associated loci in *C. difficile*.

## Introduction

Stem cell transplantation (SCT) remains a cornerstone therapy for hematologic malignancies. A critical component of SCT is the conditioning regimen administered prior to transplantation. Conditioning typically consists of high-dose chemotherapy and/or radiation designed to eradicate malignant cells, suppress host immunity, and create bone marrow space to facilitate engraftment. However, although essential for engraftment, conditioning is frequently associated with significant toxicity. Therefore, development of treatment-related adverse events often necessitates interruptions in dosing schedules or reductions in dose intensity, thereby compromising progression-free and overall survival in cancer patients. We have previously reported that approximately 40% of patients receiving cytotoxic cancer chemotherapy (CCC) develop high-grade adverse events involving the lower gastrointestinal tract.^1^ (Apewokin et al., 2015) Notably, these toxicities were not only dose-dependent but also independently associated with *Clostridioides difficile* infection (CDI). *C. difficile* remains the most prevalent healthcare-associated pathogen, and patients undergoing conditioning chemotherapy are nearly ten times more likely to develop CDI than chemotherapy-naïve individuals. It is unknown whether conditioning regimens exert direct effects on the microbe in vivo that promote CDI. The objective of this study is to address this critical knowledge gap and elucidate the interactions between conditioning chemotherapy and *C*.*difficile* that contribute to *C. difficile* pathogenesis. C. difficile has a toxin producing genetic element known as the Tcd gene region, which is composed of genes such as TcdA, TcdB, TcdC, TcdE, and TcdR genes. We specifically wanted to assess the epigenetic changes that are associated with *C*.*difficile* toxin production in the context of cancer chemotherapy exposure.

## Methods

### Patients and study design

Patients scheduled to receive a stem cell transplant preparative regimen were consented under an IRB approved protocol and provided longitudinally collected stool samples for the study that was stored at -80 degrees until ready for processing. Stool samples were collected at baseline prior to the start of the conditioning regimen and after the conditioning regimen has been started but prior to any antimicrobials. Fresh unpreserved stool samples were collected in sterile containers and frozen to –80C until culture processing could occur. Culture preservatives such as Cary Blair were not used in culturing techniques. At time of processing fresh frozen stool samples were allowed to come to room temperature prior and screened for the presence of *C. difficile* by positive detection of either Glutamate Dehydrogenase surface antigen, toxin A/B antigen, or targeted genes (Toxin B, Binary Toxin 027, and tcdC) by the use of Abbot C.diff Quik Chek. Complete and Cepheid *C*.*difficile*/epi test kits. Samples that tested positive for any C. difficile marker were subsequently processed for culturing.

### Bacterial Growth

Fresh stool was suspended in sterile saline using a sterile swab followed by vortex. 1mL of stool-saline suspension was then added, using anaerobic and aseptic techniques, to CCMB-TAL broth media (by Anaerobic Systems) to screen for the growth of C. difficile while inhibiting the growth of most fecal contaminants. CCMB-TAL broths were incubated at 37-39C for 2 days and then checked for growth. Broths negative for growth were held for an additional 5 days (for a total of 7 days) to check for delayed growth of C. difficile. Positive broth cultures were then plated onto CHOMAgar® C. difficile media and streaked for isolation to allow further selection and differentiation of *C*.*difficile* isolates from fecal microbiota. Black colonies *C. difficile* were then subcultured to TSA with Sheep Blood agar from Thermo Scientific using isolation methods to provide purified cultures. Black colonies with atypical *C*.*difficile* morphology were gram stained to confirm cellular morphology. Pure *C. difficile* isolates from TSA with Sheep Blood agar subcultures were then suspended in PBS for sequencing use. *C*.*difficile* isolates were then sequenced and underwent genome assembly for further confirmation.

### Epigenetic Profiling

We used DNA extracted with Qiagen kit then sequenced using the nanopore platform. We removed adapter sequences with porechop and then performed epigenetic profiling with the software nanodisco.5 Methylation probability was then compared between the experimental groups. Prior to epigenetic profiling we assembled the genomes of the isolates and performed phylogenetic analysis to confirm that the strains are genetically related. Genomic DNA was sequenced via Oxford Nanopore Technology nanopore sequencing and Illumina short read sequencing. A hybrid genome assembly was created for each sample via FYLE.

### Bioinformatics analysis

Genomic binning and data visualization was performed using Methylartist (https://github.com/adamewing/methylartist).^2^,^3^ Genomic bins created from Methylartist were graphed and underwent statistical testing via GraphPad Prism 10.

## Results

Assessment of the methylation profile of Tcd region revealed significant differences in methylation probabilities.

### Toxin expressing samples of C. difficile are epigenetically different from each other

In this study, a genome wide methylation profile revealed that *C. difficile* isolates displayed a diverse range of epigenetic modifications with N6 – methyladenine (6mA) being the most frequent followed by 5-methylcytosine (5mC). When isolates were grouped by toxin expression status, a significant difference in the mean 6mA methylation probabilities was observed between toxin-positive and toxin-negative strains (0.1224 vs. 0.1139, p < 0.0001, Figure 1). Epigenetic profiling across the whole genome revealed toxin-negative isolates consistently exhibited lower baseline 6mA methylation density than toxin-positive isolates across the genome.

**Figure 1.**
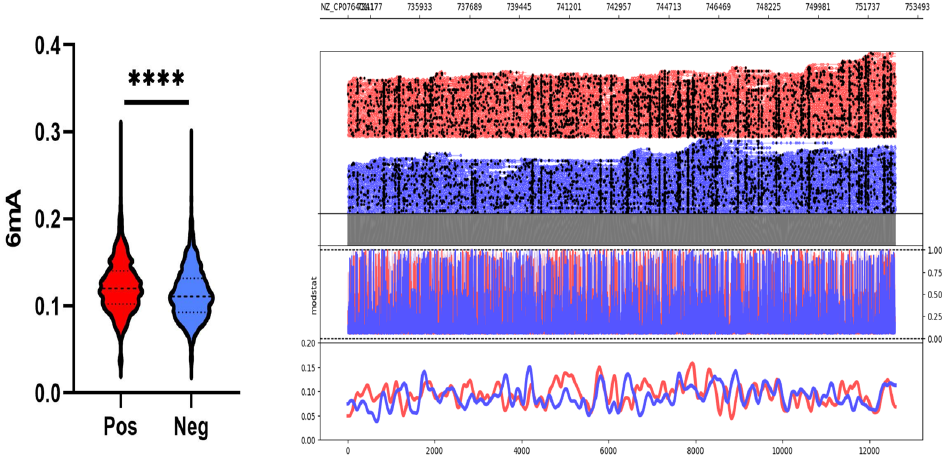
Whole genome 6mA methylation profile of C. difficile isolates grouped by positive and negative toxin expression. Probabilities were calculated by averaging predicted methylations across 200 bp genomic bins. A significant difference in 6mA methylation probability was observed between toxin positive and toxin negative isolates (0.1224 vs. 0.1139, p < 0.0001).

**Figure 2.**
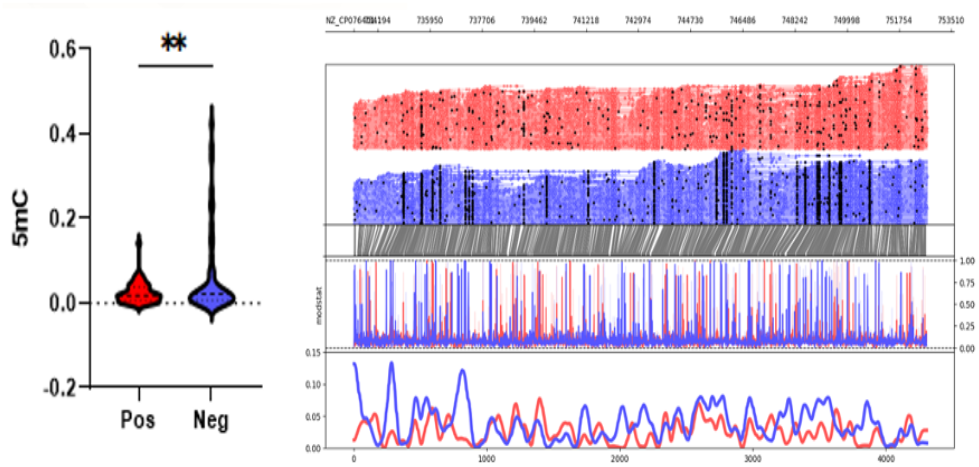
5mC methylation profile around the TcdB region (+1000 upstream and downstream) of C. difficile isolates grouped by positive and negative toxin expression (0.02565 vs. 0.06863, p = 0.0095). Top graph shows sequencing coverage while black dots indicate individual methylations. Middle graph shows the probability of methylation at each position based on modstat calculations. Bottom graph provides a smoothed representation of methylation probability across the genome.

Focused analysis of the TcdB pathogenicity locus revealed higher mean methylation probabilities for 5mC (0.02565 vs. 0.06863, p = 0.0095) but not 6mA (p = 01798) in toxin-negative samples. Methylation analysis across the TcD gene cluster, confirmed significant differences in TcdA and TcdB (adjusted p value < 0.05) but not in TcdC TcdD or TcdE.

### Conditioning chemotherapy potentially causes epigenetic changes in C. difficile

To assess whether conditioning chemotherapy impacted the epigenetics of *C. difficile*, paired isolates from patients sampled before (period A) and after (period B) were compared. For Figure 4, we saw that there was a significant difference between the number of 6mA methylations for period A and B samples (p < 0.0001). Once again, across all samples, C difficile isolates displayed a significant increase in methylated adenines compared to other methylation types as expected due to the increased role of 6mA modifications in bacterial gene expression and cell cycle.

**Figure 3.**
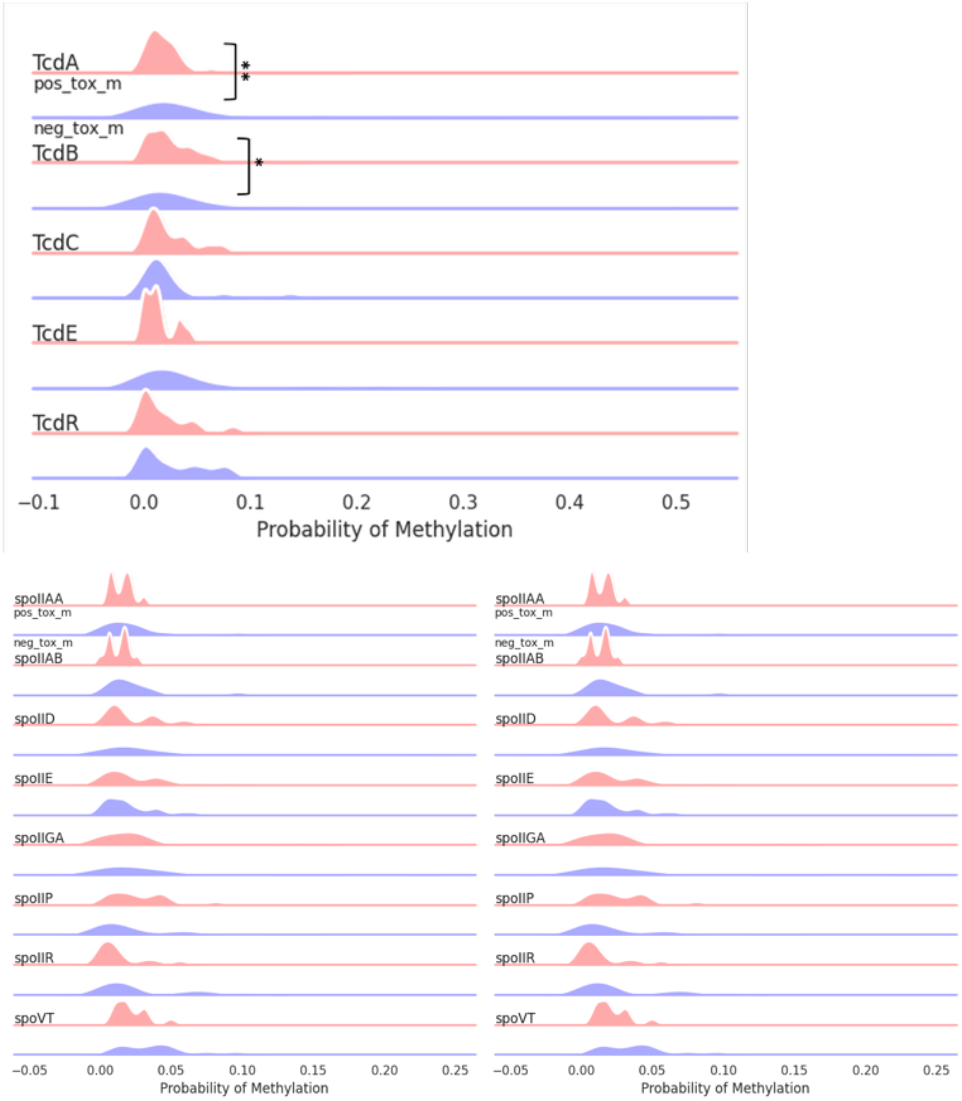
5mC methylation profiles of toxin expression groups across toxin (Tcd A,B,C,E,R) and sporulation related (spoVT, yabQ, etc.) genes. TcdA and TcdE showed statistically significant methylation changes. Some sporulation genes (spoIID, spoIIGA, spoVT) were unmethylated in toxin-negative isolates. Percentages were calculated for every 200 bp genomic bin.

**Figure 4.**
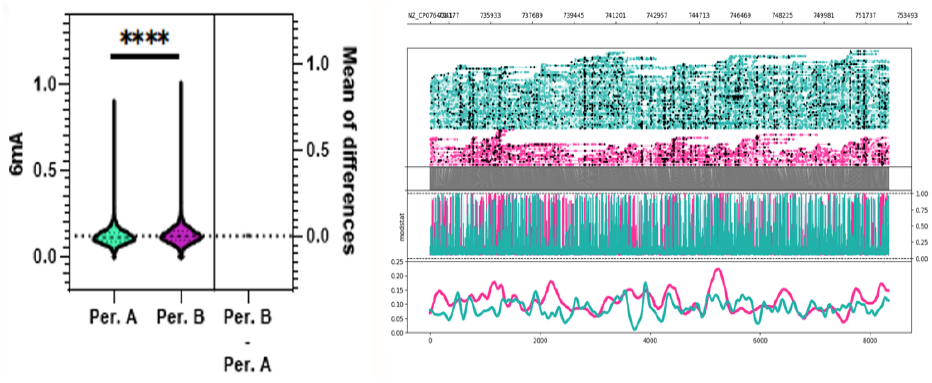
Whole genome 6mA methylation profile C. difficile isolates grouped by period A, before chemotherapy, and period B, after chemotherapy. Probabilities were calculated by averaging predicted methylations across 200 bp genomic bins. A significant difference in mean 6mA methylation probability was observed between period A and period B isolates (MD = 0.00712, p < 0.0001).

Strikingly, we noted for TcdA and TcdB of the Tcd pathogenicity locus, there was significantly lower unmethylation after chemotherapy treatment was received (**Figure 5 & 6**). p = 0.0082

**Figure 5.**
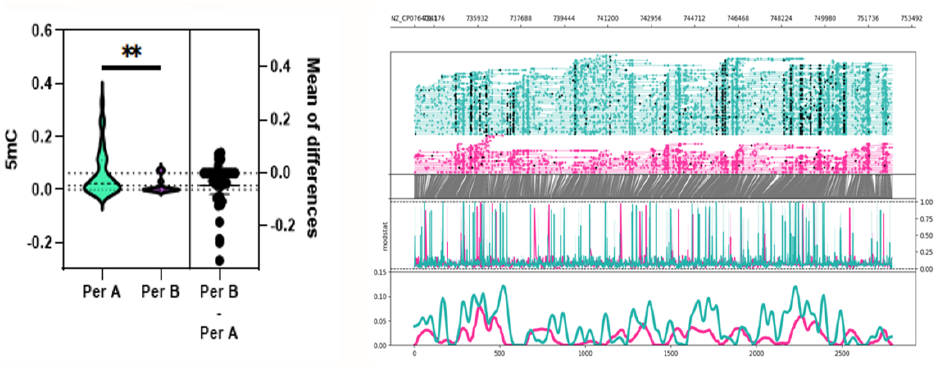
5mC methylation profile around the Tcd region (+1000 upstream and downstream) of C. difficile grouped by period A, before chemotherapy, and period B, after chemotherapy. Top graph shows sequencing coverage while black dots indicate individual methylations. Middle graph shows the probability of methylation at each position based on modstat calculations. Bottom graph provides a smoothed representation of methylation probability across the genome. A significant difference was observed for mean 5mC methylation probability around the TcdB region between period A and period B isolates (MD = 0.0464, p = 0.0082).

**Figure 6.**
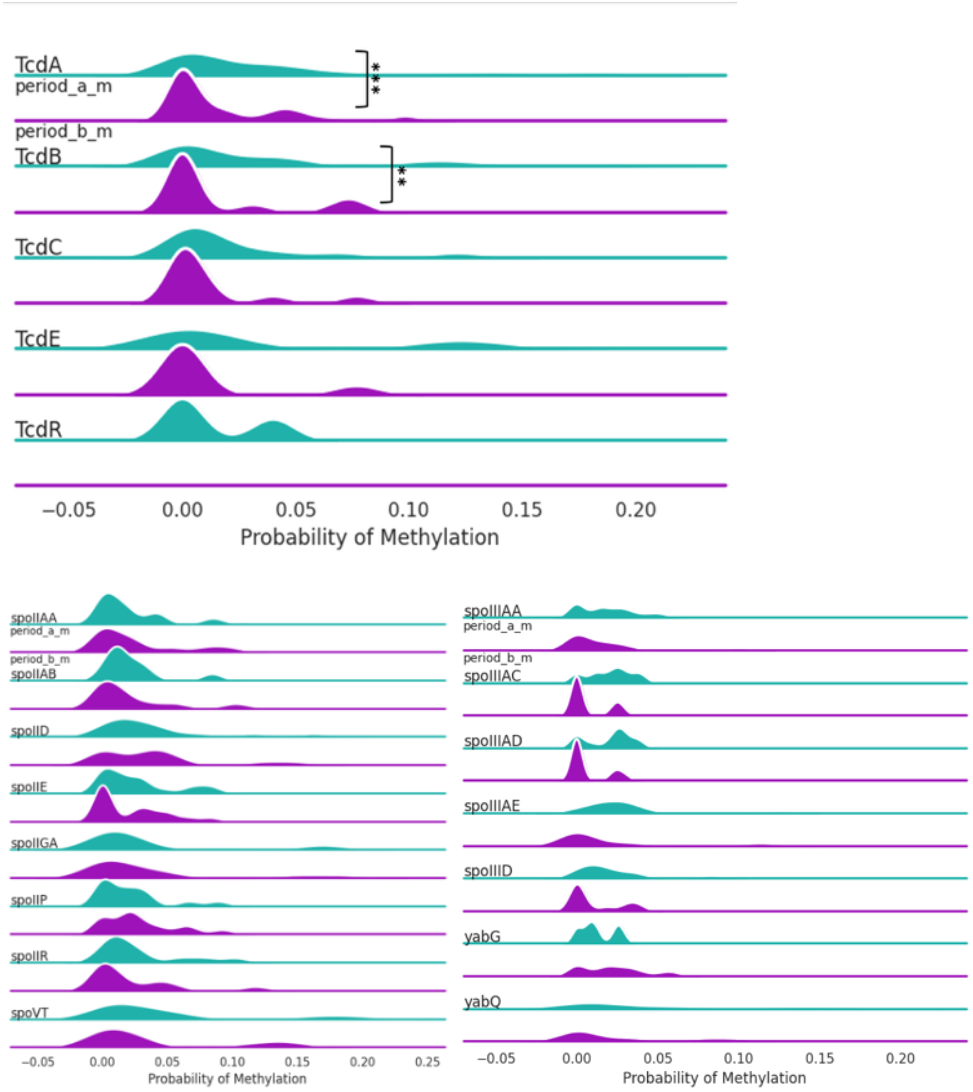
5mC methylation profiles of period A and B groups across toxin (TcdA, B, C, E, R) and sporulation related genes. In genes TcdA, TcdB, TcdC and TcdE, there were lower percentages of methylation after chemotherapy. While there was some variation, there were not any significant differences between the groups for sporulation genes. Percentages were calculated for every 200 bp genomic bin.

## Discussion

*Clostridioides difficile* is a highly methylated, spore-forming anaerobe within the gut microbiome and the etiologic agent of C. difficile infection (CDI). CDI pathogenesis is primarily mediated by the exotoxins TcdA and TcdB, which disrupt intestinal epithelial tight junctions, induce cytoskeletal disorganization, and elicit robust inflammatory responses. Chemotherapy markedly increases the risk of CDI—by nearly an order of magnitude compared with the general population— largely through disruption of the gut microbiota, mucosal barrier injury, and immune suppression. However, the effects of chemotherapy on *C. difficile* pathogen biology and toxin regulation remain incompletely understood.

Here, we provide evidence that epigenetic modifications—specifically DNA methylation of the toxin-encoding genes *tcdA* and *tcdB*—are associated with chemotherapy exposure in *C. difficile* isolates from stem cell transplant (SCT) recipients. A major strength of this study is the longitudinal sampling design, which enabled comparison of isolates obtained before and after initiation of conditioning chemotherapy, allowing evaluation of changes within the same strain. We observed statistically significant differences in methylation probabilities following chemotherapy exposure. In addition, isolates that produced toxin B exhibited distinct and statistically significant methylation patterns compared with non–toxin-producing *C. difficile* isolates. Collectively, these findings suggest that chemotherapy-associated epigenetic remodeling of *C. difficile* may contribute, in part, to the increased incidence and severity of CDI in chemotherapy-exposed patients.

Investigation of methylation patterns in bacterial pathogens has historically been limited by the technological constraints of commonly used sequencing platforms. Short-read sequencing technologies, such as Illumina, do not detect N6-methyladenine (6mA) or N4-methylcytosine (4mC), which represent major epigenetic modifications in prokaryotes. Recent advances in long-read sequencing platforms, including nanopore and PacBio, have overcome these limitations and have led to a growing body of literature examining bacterial epigenetics and its clinical relevance.

In 2015, van Eijk *et al*. reported the first comprehensive analysis of DNA methylation in *C. difficile*, identifying 6mA and 4mC modifications in the reference strain 630.^4^ Subsequently, Oliveira *et al*. provided functional insights into these methylation patterns and identified the methyltransferase *camA*, which also functions as a sporulation regulator, in 36 clinical isolates and more than 300 publicly available genomes.^5^ Notably, both studies relied on PacBio sequencing, which has limited sensitivity for 5-methylcytosine (5mC) detection compared with nanopore sequencing.

Despite these limitations, the clinical relevance of bacterial epigenetic regulation has been supported by studies repurposing epigenetic inhibitors to target *camA*, resulting in altered sporulation and biofilm formation.^6-7^ However, these studies did not include patient-level clinical data or information regarding prior drug exposures, limiting interpretation of the drivers of these epigenetic changes.

Cancer chemotherapy is a well-established risk factor for CDI, yet the mechanisms underlying this association remain poorly defined. Several chemotherapeutic agents can influence both host and microbial biology. For example, Otsuka *et al*. demonstrated that etoposide induces global DNA methylation changes in induced pluripotent stem cell (iPSC) lines and highlighted additional conditioning-regimen agents with variable epigenetic effects.

Unfortunately, there is a paucity of studies that evaluate the effects of these agents on bacterial cells. Anticancer drugs have been demonstrated to induce stable, regulatory epigenetic modifications in bacteria. Drugs such as doxorubicin and etoposide can trigger transient DNA damage responses, while fluoroquinolones can induce SOS-mediated phenotypic persistence, but these effects arise from stress-signaling and mutagenesis rather than inherited epigenetic programming. Since bacteria utilize 6mA and 4mc mechanisms which are not detected by short read sequencing platforms the paucity of data may be a reflection of this rather than an absence of biology. Intriguingly we noted an overall increase in whole genome 6mA methylation in the toxin producing strains, this may be due to increased accessibility to RNA polymerase via local structural changes and interference with repressors like CodY and CcpA. 6mA methylation could also regulate phase variation, support DNA repair fidelity and regulate metabolic shifts. *C. difficile* has a toxin producing gene region known as the Tcd gene region, which is composed of genes such as TcdA, TcdB, TcdC, TcdE, and TcdR genes. Our finding that toxin-producing isolates exhibit higher global 6mA methylation suggests that adenine methylation may function as a regulatory and genomic stabilization mechanism supporting virulence gene expression, maintenance of the PaLoc, and stress-adaptive phenotypes characteristic of pathogenic C. difficile strains. It is also particularly interesting that 5mC methylation was lower in toxin producing strains of C.diff. Many bacteria do not utilize 5mC methylation except a notable few that include, *Bacillus, Clostridia, Mycobacteria* and *Streptococcus*. In these bacteria 5mC is usually tied to restriction-modification systems and in some cases gene expression regulation. The loss of methylation in the TCD A and B region after chemotherapy exposure in the toxin producing strains therefore suggests regulatory gene modification by these agents. Understanding this mechanism may open avenues for novel interventions, such as methyltransferase inhibitors or microbiome stable medications that can prevent demethylation or potentially completely halt C. difficile epigenetic modifications.

## Limitations

Given that the *C*.*diff* had to isolated and cultured from stool, it is possible that some of the epigenetic changes may have occurred during the samples processing. However there will likely still be changes that persisted through the processing since methylation changes tend to be inherited.

## Data Availability

All data produced in the present study are available upon reasonable request to the authors

## Conclusions

*C. difficile* exhibits changes in epigenetic profiles that is associated with chemotherapy exposure and correlates with toxin production. These changes may explain the increased incidence of *C*.*diff* observed in chemotherapy exposed patients.

